# The eSAH Score: A Simple Practical Predictive Model for SAH Mortality & Outcomes

**DOI:** 10.1101/2023.09.15.23295634

**Authors:** Rohan Sharma, Daniel Mandl, Fabian Foettinger, Saif Salman, Raja Godasi, Yujia Wei, Rabih Tawk, W David Freeman

## Abstract

**Background:** We developed a simple quantifiable scoring system that predicts aneurysmal subarachnoid hemorrhage (aSAH) mortality, delayed cerebral ischemia (DCI) and modified Rankin Scale outcomes using readily available SAH admission clinical data with a new radiographic quantitative volumetric SAH method.

**Methods:** We analyzed 277 patients with aneurysmal SAH (aSAH) admitted at our Comprehensive Stroke Center (CSC) at Mayo Clinic Florida between 2012 and 2022. We developed a mathematical model that measures aSAH basal cisternal subarachnoid hemorrhage volume (SAHV) using a derivation of the ABC/2 ellipsoid formula, where A = width/thickness, B = length, C = vertical extension) on non-contrast CT (NCCT), which we previously demonstrated comparable to pixel based manual segmentation on NCCT scans. Data was analyzed using t-test, chi-square, receiver operator characteristics (ROC) curve, and area under curve analysis. Multivariate logistic regression analysis with stepwise elimination of variables not contributing to the model (0.05 significance level for entry into the model) was used to develop an enhanced SAH (eSAH) scoring system.

**Results:** Using regression and logistic regression, we found that age, GCS score and SAHV were significantly associated with final discharge outcome, prediction on in-hospital DCI, and in-hospital mortality. A weighted eSAH score was developed using these factors that ranged between ‘0-5’ and was strongly predictive of outcome (AUC=0.88), DCI (AUC=0.75) and in-hospital mortality (AUC=0.87).

**Conclusions:** A volumetrically-enhanced SAH (eSAH) score is a simple quantitative model based on SAH volumetrics, GCS and age and appears to predict mortality and outcomes in SAH patients. A larger cohort validation study is planned.

## Introduction

Aneurysmal subarachnoid hemorrhage (SAH) is a devastating hemorrhagic stroke subtype that occurs in about 30,000 patients per year in the United States and is with an estimated 30-40% one-month historical mortality.(1-3) The high morbidity and mortality of SAH are due to both primary neurological injury as well as a cascade of secondary neurological injury that ensues from inflammation cascade in response to blood in the subarachnoid space, including cerebral vasospasm, delayed cerebral ischemia (DCI), hydrocephalus.(4-6) Although several SAH grading and scoring systems have been proposed to predict outcomes for aSAH, they have limited predictive capability due to an imprecise, semi-quantitative Fisher scale measurements of SAH blood volume (7-13). Our study aimed at developing a simplified predictive model based on admission clinical and radiological features at initial presentation to predict outcomes for aSAH and using a new quantitative SAH <u>v</u>olumetric (SAHV) measurement scale using a modified ABC/2 derived methodology. Despite decades of research there remains only 1 FDA-approved drug nimodipine approved for neuroprotection in SAH patients(14). We feel that accurate measurement of SAHV blood volume is critical for future SAH discovery, translation and drug application for new drug targets and interventional drug trials.

## Methods

### Study Cohort

We conducted retrospective analysis of cohort of 277 patients in the electronic medical record of Mayo Clinic (Jacksonville, Florida) with a diagnosis of SAH admitted at the Mayo Clinic Emergency Medical Department between 2012 and 2022. Demographics, past medical history and preexisting conditions, clinical assessment, and CT imaging studies at admission as well as clinical management and course during hospitalization were retrospectively acquired via electronic medical record database EPIC. The data collected was stored in two separate electronic databases in RedCap. Neurological deterioration due to DCI was derived from electronic medical record notes, progress reports and neurological imaging and defined according to Vergouwen et. al. [10] Based on clinical assessments at discharge modified Rankin Scale (mRS) could be abstracted according to the Specification Manual for Joint Commission National Quality Measures (v2018A).

#### Baseline Characteristics

Several criteria had to be met to be included in this study. Initial non-contrast computed tomography imaging (NCCT) and SAH diagnosis had to be done within 24 hours of SAH ictus. The CT imaging data had to have 5 mm imaging slices or less or at least equivalent reformatting to measure the data quantitatively. Electronic medical record of clinical course had to be present as defined by standard of care (SOC). Neurological imaging had to be present to definitively diagnose and further relativize the extent of DCI, including computed tomography angiography (CTA), magnetic resonance tomography (MRT), magnetic resonance angiography (MRA) or digital subtraction angiography (DSA). Functional outcome at discharge days had to be extractable/abstractable from the electronic medical record and was graded according to modified mRS.

Definitive criteria for exclusion were absent CT imaging, no diagnosis of SAH or associated disease (only intracerebral hemorrhage, IVH or subdural hemorrhage without SAH) and traumatic SAH defined by major traumatic history and no record of ruptured aneurysm. Patients who died within 72 hours of admission had to be excluded from DCI and sVSP analysis, as the time of onset for these secondary complications typically exceeds this time frame.

#### Imaging Analysis and SAHV Methodology

All admission NCCT were stored either as Digital Imaging Communications in Medicine format (DICOM) or Neuroimaging Informatics Technology Initiative (NIfTI) format. Imaging analysis was performed on a personal computer using Mayo Clinic medical imaging software “QReads” for determination of metric extensions (Version 5.14.0, Mayo Clinic).

Estimated volumetric data on non-contrast CT images were analyzed using a quantitative SAH volumetric approach (SAHV). We have reported the methodology and cisternal anatomic definitions a prior publication (15). For this method, cisternal spaces were predefined and by assuming that the basal cisterns morphology exhibits an ellipsoid configuration, cylindrical volumetrics were estimated by calculating volume through application of ABC/2 (with A = width/thickness, B = length, C = vertical extend), and hemorrhagic volumes thereby measured. This methodology has been established for calculating intracerebral hemorrhage (ICH) volume(16). The subsequently estimated volumes were then summed to a total cisternal hemorrhagic blood volume (SAHV). We have previously reported this method to be comparable to voxel based manual segmentation method.

#### Outcomes Assessment

Discharge modified Rankin scale (mRS) was used as outcome data and was dichotomized with mRS 0-3 as favorable outcome and 4-6 as unfavorable outcome. For univariate analyses, independent variables were compared using χ2, Student t-test (as appropriate). The outcome model was developed using multivariate logarithmic regression analysis with all possible prediction variables that would be available at the time of initial presentation (including gender, age, SAH volume, Glasgow Coma Scale (GCS), modified Fisher’s score (mFS), Hunt and Hess scale, presence of intraparenchymal hemorrhage, presence of intraventricular hemorrhage). The analysis was then followed by stepwise elimination of variables not contributing to the model (0.05 significance level for entry into the model). First order interactions were tested in the final model.

An outcome stratification model for volumetrically enhanced Subarachnoid Hemorrhage Score called eSAH score was created with the variables in the final outcome model. Cut off points of variables were chosen to produce a simple and intuitive model. A DCI subscore was similarly calculated for risk stratification model for DCI prediction based on labeled DCI outcomes in the dataset using the consensus DCI criteria of Vergouwen et al(17). Nonparametric two-Sample Kolmogorov-Smirnov Test was performed to test the distribution of outcome and in-hospital mortality with eSAH score, and DCI with DCI subscore. Discriminative accuracy of the score was examined using receiver operator characteristics curve and subsequent area under the curve analysis. Analyses was done using SAS software version 9.4. Logistic regression models were built in SAS and from this calculation of the odds ratio by exponentiating the log odds.

## Results

Out of 277 patients with SAH, 72 were deemed to be traumatic in nature and were excluded from analysis. Demographic data about 205 non-traumatic SAH including age, sex, admission GCS etc. have previously been reported(15). We excluded additional 15 patients who had hemorrhage only in extracisternal spaces but no hemorrhage in described cisternal spaces were also excluded from analysis, leaving 190 SAH patients for analysis. Overall 30 day mortality rate in this cohort was 15.79%. The mean cisternal hemorrhagic volume in this corhort was 11.65 mL (range 0.13 mL to 116.9 mL).

Outcome at discharge was found to be statistically associated with age (p<0.001), SAH cisternal volume (p<0.001), GCS (p<0.001), mFS (p<0.001), Hunt and Hess score (p<0.001), presence of intraparenchymal hemorrhage (p=0.02) and intraventricular hemorrhage (p<0.001) in univariate analysis. Gender was not significantly associated with outcome (p=0.08).Out of all variables found to be correlated to outcome in univariate analysis, on conducting the stepwise multivariate regression analysis, only cisternal hemorrhagic volume, age and GCS were found to statistically contribute to the overall outcome. On conducting a similar analysis for DCI, only GCS and cisternal hemorrhagic volume were found to significantly contribute to the model.

Outcome risk stratification was developed for all nontraumatic SAH patients with aim of developing a simplified predictive model. Age, GCS and SAH cisternal volume were predictive of outcome at the time of discharge as well as in-hospital mortality (Table 2). For every increase in SAHV by 1 ml, the odds of unfavorable outcome increased by a factor of 1.148 (95% confidence interval (CI) = 1.973 – 1.227). For every increase in age by 1 year, the odds of unfavorable outcome increased by a factor of 1.051 (95% CI = 1.015 – 1.087). For every increase in GCS by 1 point, the odds of unfavorable outcome decreased by a factor of 0.728 (95% CI = 0.651 – 0.815). Subsequently, the eSAH score was created with cutoffs in age, GCS and SAH cisternal hemorrhagic volume to create a simple risk stratification tool for prediction outcome and mortality. Given that DCI was only associated with two variables GCS and SAHV, the eSAH DCI subscore was calculated and derived with these variables of GCS and SAHV.

Cut offs were made to change the variable to an ordinal scale for development of scoring system and the point assignment for each variable is described in **Table 1**. The eSAH score therefore was calculated as a summation of individual points for each variable. The eSAH score ranged from 0 to 5 and the eSAH DCI subscore ranged from 0 to 4 for subsequent risk of developing DCI. The eSAH score at admission was strongly predictive of outcome at the time of discharge with increase in odds of poor outcome by a factor of 4 for increase in every point for eSAH score{OR= 4.27 (95% CI=2.84 – 6.81), p<0.0001, AUC=0.885). It was also a strong predictor of in-hospital mortality with threefold increase in odds of mortality with every point increase in eSAH score{OR=3.02 (95% CI = 1.98 – 4.61), p<0.001, AUC=0.878). The eSAH DCI subscore was also strongly predictive of DCI development and had doubling of odds of DCI with point increase in DCI subscore {OR = 1.97 (95% CI = 1.49 – 2.59), p=0.001, AUC=0.748).

**Table 1:** Enhanced SAH (eSAH) Score eSAH score is calculated by scoring each category points allotted by variable and summing them up. Total eSAH score = GCS score + Age score + SAH Volume (SAHV) score. SAHV is calculated based on work of Fottinger et al(15). using a simplified ABC/2-derived method or automated method, minimum eSAH score = 0, Maximum eSAH score= 5. DCI risk subscore = GCS score + SAH volume score, Minimum score = 0, Maximum score= 4

| **Points** | **Variable** |
| --- | --- |
| **Glasgow Coma Scale (GCS) Points** | **GCS** |
| **0** | 12-15 |
| **1** | 8-11 |
| **2** | 3-7 |
| **SAH Volume (SAHV)\*** | **SAHV** |
| **0** | Less than 10 ml |
| **1** | 10-20 ml |
| **2** | Greater than 20 ml |
| **Age** | **Age** |
| **0** | Less than equal to 60 |
| **1** | Greater than 60 |

**Table 2:**

| Multivariate Regression Analysis of Outcome in SAH patients |  |  |  |  |  |  |
| --- | --- | --- | --- | --- | --- | --- |
| Variable | Odds Ratio (95% confidence interval) |  |  |  |  |  |
| Age | 1.051 (1.015 – 1.087) |  |  |  |  |  |
| SAHV | 1.148 (1.073 – 1.227) |  |  |  |  |  |
| GCS | 0.728 (0.651 – 0.815) |  |  |  |  |  |
| Distribution of outcome by eSAH score |  |  |  |  |  |  |
|  | eSAH score |  |  |  |  |  |
| Outcome | 0 | 1 | 2 | 3 | 4 | 5 |
| Favorable | 51 (92.73%) | 36 (75%) | 16 (48.48%) | 2 (6.45%) | 1 (7.69%) | 0 |
| Unfavorable | 4 (7.27%) | 12 (25%) | 17 (51.52%) | 29 (93.55%) | 12 (92.31%) | 10 (100%) |
| Distribution of in-hospital mortality by eSAH score |  |  |  |  |  |  |
|  | eSAH Score |  |  |  |  |  |
| Death | 0 | 1 | 2 | 3 | 4 | 5 |
| No | 55 (100%) | 47 (97.92%) | 28 (684.85%) | 20 (64.52%) | 6 (46.15%) | 4 (40%) |
| Yes | 0 | 1 (2.08%) | 5 (15.15%) | 11 (35.48) | 7 (53.85) | 6 (60%) |
| Distribution of DCI by DCI subscore |  |  |  |  |  |  |
|  | DCI subscore |  |  |  |  |  |
| DCI | 0 | 1 | 2 | 3 | 4 |  |
| No | 63 (92.65%) | 35 (171.43%) | 18 (62.07%) | 8 (42.11%) | 7 (46.67%) |  |
| Yes | 5 (7.35%) | 14 (28.57%) | 11 (37.93%) | 11(57.89%) | 8 (53.33%) |  |

Outcome, in-hospital mortality and DCI Data

Among the 50 patients with an eSAH score of 0, zero died and 46 (92%) had favorable outcomes. In contrast, out of 12 patients with score of 5, 0 had a favorable outcome and 7 (58%) ended up dying during the hospital stay. Out of 68 patients with DCI subscore of 0, only 5 (7%) developed DCI as opposed to greater than 50 percent of patients with DCI subscore of 3 & 4.

## Discussion

Clinical severity scoring systems have been used in triaging and guiding interventions and treatment for several illnesses. Examples of such scoring systems include GCS, NIH Stroke Scale, mFS, Hunt and Hess scale, Intracerebral hemorrhage (ICH) score, Sequential Organ Failure Assessment (SOFA), Clinical Institute Withdrawal Assessment for Alcohol (CIWA) etc. and many of these are used widely in clinical care(18-24). Since the proposition of C. Miller Fisher’s original (21, 22) SAH scale and its subsequent modification by Frontera et. al. (25), no other widely used radiologic model has been established to quantify hemorrhagic volumes in aSAH and risk for poor outcome and DCI, due to lack of clinical applicability or prognostic value (7, 26, 27). Other clinical scores such as Hunt and Hess scale, the world federation of neurosurgeons scale (WFNS), SAH score have also been developed to assess for surgical risk or mortality but do not have radiological input and have limited clinical utility (28-30). There is a need for a grading system for SAH that is objective and imputes SAH severity from both clinical and radiological information and can serve as a robust predictive tool for outcome. ICH score is one such clinico-radiologic score that predicts mortality of patients with ICH (18).

Previous studies found a strong association between subarachnoid hemorrhage volume, clot thickness, hemorrhagic persistence, and concomitant intraventricular hemorrhage with DCI and functional outcome(5, 31). We used a simplified (ABC/2-derived) SAHV method and manual segmentation to conduct volumetric analysis of SAH with in the cisternal spaces. We have previously reported that it yielded similar results in terms of volume estimation when compared to voxel based manual segmentation analyses. Volume of hemorrhage has been found to correlate with outcome in ICH, and it is axiomatically assumed that it would have similar effects on SAH outcome. The modified Fisher scale however is only a semiquantitative method of measuring SAH blood and is not as reliable as direct volumetric analysis for prediction of outcome and DCI(15) (). Our previous work had already demonstrated a direct association of SAH volume with outcomes and DCI. We subsequently aimed to incorporate additional clinical covariates and found that age and GCS expounded the prediction capability of SAHV in terms of outcome at discharge and in-hospital mortality. The discriminative accuracy of this score for both clinical outcome and death was found to be robust. Also, the DCI subscore which included only GCS and SAHV was found to be fairly predictive of future DCI that occurs after SAH.

In order for a score to be clinically useful in variable settings, it has to be simple and reproducible similar to the ICH score by Hemphill such that others can validate. The eSAH score is simple and easy to use with only three parameters. Two of the three variables include age and GCS should be readily available in low resource critical access hospitals and emergency departments and different clinical settings such as primary stroke centers and comprehensive stroke centers. One limitation of this model is diffusing knowledge of our ABC/2 derived SAH volumetric (SAHV) scale. However, the ABC/2 volume for intracerebral hemorrhage eventually became widespread and used and incorporated into Hemphill’s ICH score. The simplified ABC/2-derived SAHV measurement is easier to measure than manual computerized segmentation which takes considerable time. Therefore, we hope with the use of our simplified SAHV model this adds important prognostic and severity of illness information in the eSAH score. The eSAH SAVH can also be calculated like ABC/2 for ICH volume in any hospital that has non-contrast head CT scan data available and without any specialized training or imaging software.

There are several limitations to the eSAH score. First, we did not adjust for preexisting comorbidities, such as follow up imaging and clinical factors later during hospitalization such as hypertension. However, the power of the eSAH score is in its simplicity since in practice SAH patients may present without any known comorbidity information similar to the ICH score by Hemphill. Therefore, the eSAH score is best utilized with this limitation at SAH admission using only 3 variables.

Further, the eSAH score has great potential to triage and risk stratify SAH patients similar to Hemphill’s ICH score for ICH patient mortality and stroke systems of care for these patients. While we acknowledge the predictive modeling benefit of the eSAH score is limited to the 3 variables analyzed at the time of initial SAH presentation, it can be leveraged as a relative strength given the novel quantitative SAHV score which measures a “dose-response” relationship of blood volume compared to GCS with age. The SAHV is a more precise way to quantify in milliliters (ml) the amount of blood compared to the older, Likert-like modified Fisher scale. We acknowledge that both SAHV and eSAH score require further validation in a larger prospective acquired dataset to ensure its clinical utility and prognostic utility. However, the eSAH score data could be used as a risk stratification and SAH severity tool that could aid in the decision-making processes in tandem with clinician judgement.

The eSAH score could also aid the triage and transport of SAH patients from primary stroke center hospitals to comprehensive stroke center hospitals that have dedicated neuro-intensive care unit for complex SAH management as defined by the recent AHA SAH guideline(32). Patients with higher scores have a high likelihood to develop DCI and worse downstream outcomes without the multidisciplinary team that complex SAH patients require in terms of vasospasm monitoring, neuroendovascular interventions for symptomatic vasospasm and NSICU level neuromonitoring. Therefore, the eSAH score could be used in emergency departments similar to the ICH score by Hemphill to stage and document severity during the initial SAH presentation. Such eSAH score triage in the emergency department could lead to expeditious transfer to a higher-level stroke center with dedicated neurosurgical vascular and neuro-intensive care unit teams for SAH management. Alternatively, those with lower scores could be initially managed and stabilized in the nearest local stroke center. The eSAH score could therefore help achieves a more equitable allocation of stroke-center resources among stroke networks of care and as recommended by the current AHA SAH guidelines(32). The eSAH score could also potentially benefit future translational research and targeted interventions (e.g., neuroprotective drugs or minimally invasive approaches such as intraventricular calcium channel blocker drug injection) for SAH patients.

## Conclusion

The eSAH score is a simple, quantitative model based on a new quantitative SAH volumetric scale (SAHV) that incorporates readily available factors of GCS and age in the emergency department and can be used to provide estimates of SAH mortality and downstream outcomes for SAH patients. The eSAH score can also potentially help triage SAH patients at presentation to help in the acute decision-making in management of SAH patients to higher level comprehensive stroke centers with neurointensive care units and vascular neurosurgeons.

**Figure 1:**
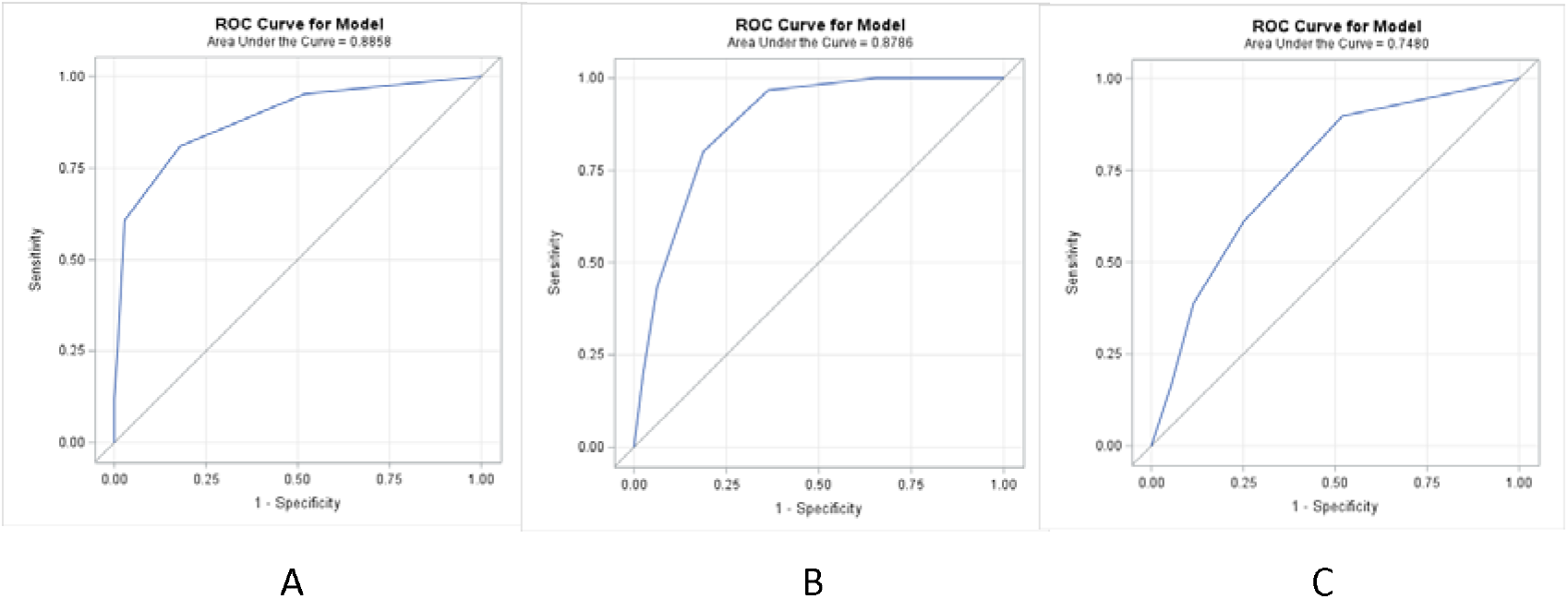
Receiver-Operating Characteristics (ROC) curves compared to various outcome measures in SAH patients in A, B, and C. A,B: ROC curve for Outcome and in-hospital mortality based on eSAH score C: ROC curve for DCI based on DCI subscore

**Figure 2:**
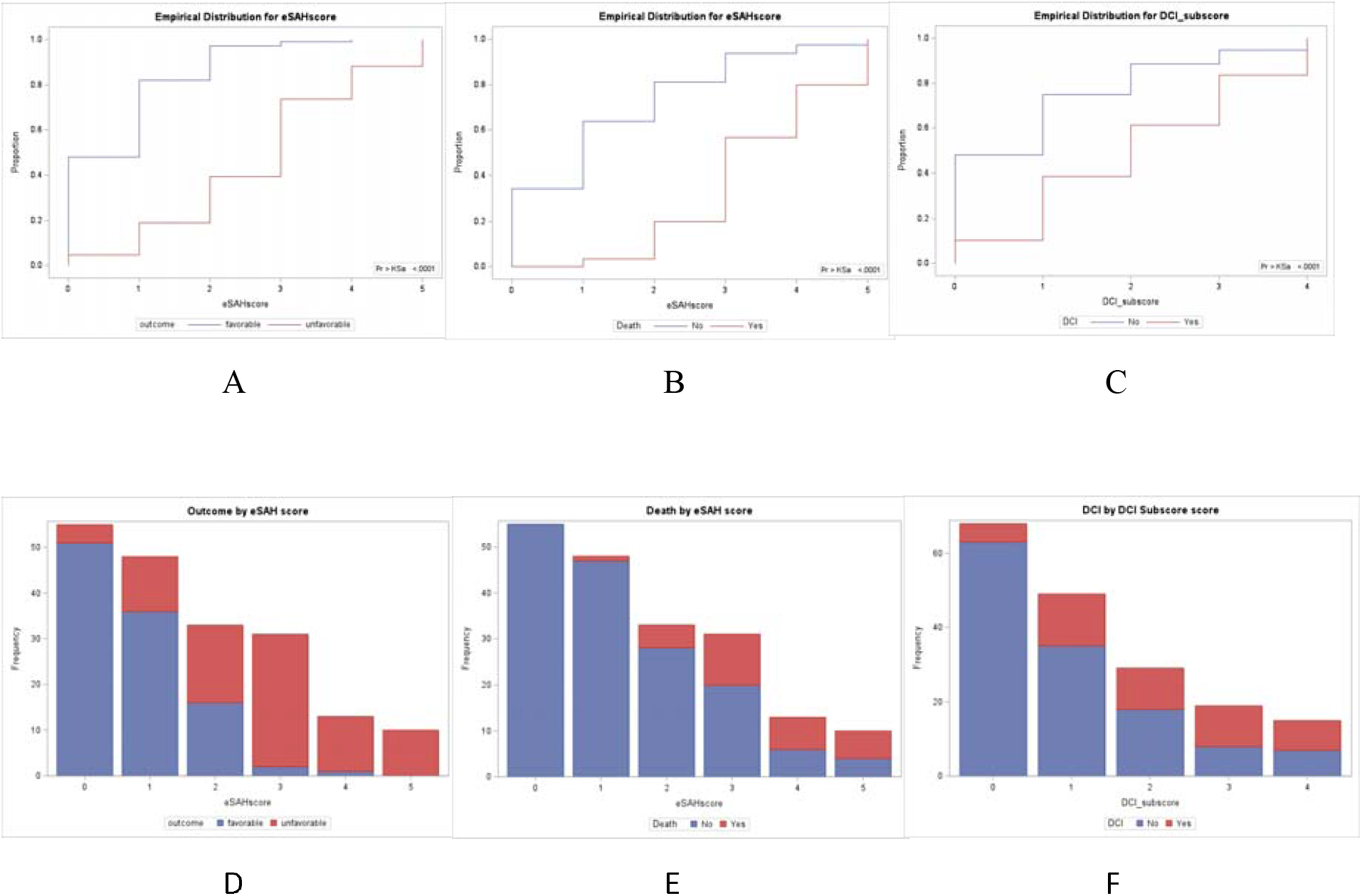
eSAH score totals with associated outcomes of mortality and DCI. A,B: Distribution of outcome, in-hospital mortality, by eSAH score. C: Distribution of DCI during admission by DCI subscore D, E: Proportion of outcome and in-hospital mortality by eSAH score F: Proportion of DCI by DCI subscore

## Data Availability

All data produced in the present work are contained in the manuscript

